# Association between longitudinal changes in left ventricular structure and function and 24-hour urinary free cortisol in essential hypertension

**DOI:** 10.1101/2024.10.04.24314927

**Authors:** Gao-Zhen Cao, Jia-Yi Huang, Qing-Shan Lin, Run Wang, Min Wu, Cong Chen, Jian-Cheng Xiu, Kai-Hang Yiu

**Author notes:** **Contact Information for Corresponding Author:** Kai-Hang Yiu, Cardiology Division, Department of Medicine, The University of Hong Kong, Hong Kong, China.; Jian-Cheng Xiu, Cardiology Division, Department of Medicine, Nanfang Hospital, Guangzhou, China.

## Abstract

**Objective:** This study aimed to examine the relationships between 24-hour urinary cortisol levels (24h-UFC) and alterations in left ventricular (LV) structure and function in patients with essential hypertension. **Methods:** A prospective cohort study was conducted at the Hypertension Center of the University of Hong Kong-Shenzhen Hospital, including 315 patients with essential hypertension. Baseline 24h-UFC levels were measured, and echocardiographic assessments were performed to evaluate left ventricular mass (LVM), left ventricular ejection fraction (LVEF), and the E/e’ ratio (early diastolic trans-mitral flow velocity to early diastolic mitral annular velocity). Patients were divided into tertiles based on their 24h-UFC levels for comparative analysis. Statistical analyses were employed to examine the relationships between UFC levels and changes in cardiac parameters over the follow-up period. **Results:** Higher baseline 24h-UFC levels were significantly associated with greater increases in LVM and E/e’ during follow-up, indicating adverse LV remodeling and diastolic dysfunction. This correlation remained significant after adjusting for confounding factors such as age, gender, baseline systolic and diastolic blood pressure, heart rate, and their changes. Patients in the highest 24h-UFC tertile exhibited an increase in left ventricular hypertrophy (LVH) prevalence, whereas those in the lower tertiles showed a reduction. **Conclusion:** Elevated 24h-UFC levels are independently associated with adverse changes in LV structure and diastolic function in patients with essential hypertension.

**Novelty and Relevance:** *What Is New?:* This is the first study to elucidate the association between longitudinal changes in left ventricular structure and function and 24-hour urinary free cortisol (24h-UFC) in essential hypertension.

*What Is Relevant?:* This cohort study indicates that higher baseline 24h-UFC levels are significantly associated with greater increases in left ventricular mass (LVM) and the E/e’ ratio during the follow-up period, suggesting adverse left ventricular (LV) remodeling and diastolic dysfunction.

*Clinical/Pathophysiological Implications?:* This study provides novel insights into the role of cortisol in cardiovascular remodeling in hypertensive patients and may have significant implications for understanding the mechanisms underlying hypertensive heart disease.

## 1. Introduction

Left ventricular hypertrophy (LVH) is a well-established risk factor for cardiovascular events including heart failure,^1^ ischemic heart disease,^2^ atrial fibrillation,^3^ and sudden cardiac death^4^ in hypertensive patients. Although robust evidence indicates that increased cardiac workload is fundamental in the development of LVH in hypertension, numerous additional non-hemodynamic factors may also be involved. These include gender, genotype, body mass, and multiple biochemical and hormonal mediators such as angiotensin II, catecholamines, steroid hormones, insulin, and some growth factors and cytokines.^5^ Additionally, LVH often occurs in conjunction with changes in left ventricular (LV) filling characteristics that reflect LV diastolic dysfunction. LV diastolic dysfunction has been shown to be associated with development of heart failure and is predictive of all-cause mortality.^6,7^ Therefore, timely identification of factors that contribute to the development of LVH and LV diastolic dysfunction is crucial to preventing functional cardiac deterioration leading to heart failure.

Hypertension is a prominent feature among patients with abnormal increases in circulating glucocorticoid levels, and cardiovascular events are a leading cause of morbidity and mortality in these patients. Previous studies conducted on patients with Cushing’s syndrome consistently demonstrate that chronic hypercortisolism results in adverse cardiac structural and functional changes that cannot be explained solely by blood pressure elevation.^8–10^ Recent research found that midnight and dexamethasone suppression test (DST) plasma cortisol levels are independent predictors of LV mass and geometry in patients with essential hypertension^11^.However, no studies have yet established a clear relationship between cortisol levels and changes in LV structure and function in patients with essential hypertension. This study aimed to examine the relationships between 24h-UFC levels and alterations in LV structure and function in patients with essential hypertension through a prospective cohort study.

## 2. Methods

### 2.1 Study design and study population

This prospective cohort study was conducted at the Hypertension Center of the University of Hong Kong - Shenzhen Hospital. A total of 414 patients diagnosed with essential hypertension at the hypertension center between July 2018 and December 2021 were included. Blood pressure measurements and hypertension diagnosis were conducted following current guidelines^12^. All patients were excluded from having secondary causes of hypertension. Cushing’s syndrome was excluded based on the measurements of plasma cortisol levels (8 am, 4 pm, 12 am), 24h-UFC, and plasma cortisol level after an overnight DST with 1 mg dexamethasone, according to the guidelines.^13^ Adrenal magnetic resonance or computed tomography imaging excluded the presence of adrenal incidentalomas. Patients with a history of ischemic heart disease (n=25), cardiac valve disease (n=11), stroke (n=11), estimated glomerular filtration rate (eGFR) <30 mL/(min·1.73 m²) (n=14), alcohol abuse (n=2), or acute infection (n=2), were excluded. A total of 349 patients were initially recruited for baseline evaluation. Patients were subsequently excluded if they developed a history of myocardial infarction (n=2), underwent percutaneous coronary intervention (n=6), or died (n=1) during follow-up. Additionally, 25 patients were excluded due to loss to follow-up. Ultimately, 315 patients were included in the final analysis (**Figure 1**). The study was conducted in accordance with the principles of the Declaration of Helsinki and received approval from the local Institutional Review Board. Informed consent was obtained from all participants.

**Figure 1.**
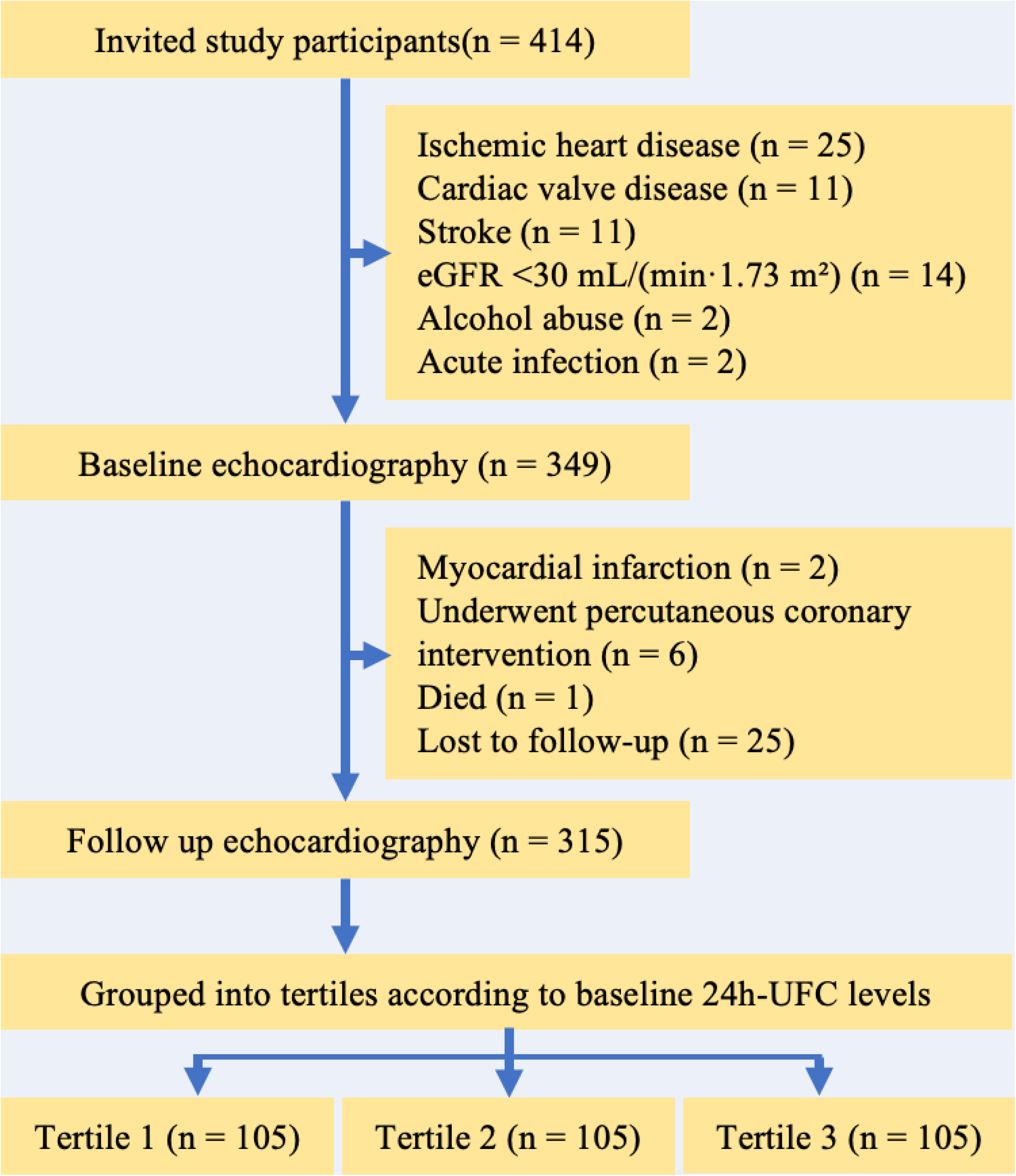
The flowchart for participants in this study.

### 2.2 Blood pressure measurement

Blood pressure was measured in accordance with the European Society of Hypertension and the European Society of Cardiology (ESH/ESC) guidelines^12^ using a validated device (Omron HEM-7130; Omron health care, Kyoto, Japan). Patients were seated comfortably in a quiet setting for at least five minutes, with their arm positioned at heart level. Initial measurements were taken from both arms, followed by two additional measurements from the arm with the highest initial reading, at two-minute intervals. The average of the final two measurements was recorded for analysis.

### 2.3 Laboratory measurements

24h-UFC levels were measured using the UniCel™ DxI 800 Access Immunoassay System (Beckman Coulter Inc., Brea, CA, USA). Morning blood samples, collected after a minimum of 12 hours of fasting, were analyzed for various biochemical parameters including glycosylated hemoglobin (HbA1c), triglycerides (TGs), total cholesterol (TC), low-density lipoprotein cholesterol (LDL), high-density lipoprotein cholesterol (HDL), serum creatinine (SCr), and adrenocorticotropic hormone (ACTH) using the Roche COBAS 8000 platform (Roche Diagnostic, Basel, Switzerland). The estimated glomerular filtration rate (eGFR) was calculated using the Modification of Diet in Renal Disease (MDRD) equation.^14^

### 2.4 Echocardiography

Standard 2D echocardiography and tissue Doppler imaging were performed using a VingmedE9 echocardiography system (General Electric Vingmed Ultrasound, Horten, Norway) by experienced operators who were blinded to the patients’ clinical and biochemical data. Images were captured in the lateral decubitus position with a 3.5-MHz transducer and stored digitally in cine-loop format. Left ventricular ejection fraction (LVEF) was calculated using the modified Simpson’s biplane method from apical 4- and 2-chamber views. Measurements of left ventricular end-diastolic diameter (LVEDd), left ventricular end-systolic diameter (LVESd), interventricular septal thickness (IVSd), and posterior wall thickness (PWTd) were obtained from 2D-guided M-mode tracings in the parasternal long-axis view, using the leading-edge method.^15^ Left ventricular mass index (LVMi) was estimated using the corrected American Society of Echocardiography (ASE) formula^15^ (0.8 × (1.04 × ((IVSd + LVEDd + PWTd)^3^ − (LVEDd)^3^)) + 0.6) and normalized to body surface area (BSA). LVH was defined as increased LVMi (≥95 g/m^2^ in females; ≥115 g/m^2^ in males). Relative wall thickness (RWT) was calculated by the ASE formula: RWT = 2 × PWTd/LVEDd, with increased RWT defined as >0.42.^15^ LV structural patterns were classified as normal (normal LVMi and RWT), concentric remodeling (normal LVMi, increased RWT), eccentric hypertrophy (LVH, normal RWT), and concentric hypertrophy (LVH, increased RWT). Left atrial volume (LAV) was quantified by tracing the endocardial borders and height of the left atrium in the apical four-chamber view, with the LAV index (LAVi) calculated by normalizing to body surface area. Peak tricuspid regurgitation (TR) velocity was measured via continuous-wave Doppler. LV diastolic function was evaluated through pulsed-wave Doppler of mitral valve inflow, measuring peak velocities during early diastole (E-wave) and late diastole (A-wave), which were used to calculate the E/A ratio. Tissue Doppler imaging measured peak early diastolic mitral annular velocity (e’) at the septal and lateral annuli, and the E/e’ ratio was calculated. Diastolic dysfunction was graded according to the 2016 ASE/EACVI guidelines using a two-step process.^16^ The first step involved assessing septal or lateral e’ velocity, E/e’ ratio, LAVi, and peak TR velocity to diagnose diastolic dysfunction. In the second step, patients with diastolic dysfunction were further graded based on the E/A ratio and E velocity followed by LAVi, E/e’ ratio, and peak TR velocity. The median follow-up period was 26 months (range 15–41 months).

### 2.5 Statistical analysis

Patients were categorized into three groups according to their 24h-UFC tertile at enrollment, with demographic and clinical characteristics analyzed separately. The Kolmogorov‒Smirnov test was used to assess the distribution of continuous variables. Normally distributed variables are presented as means ± standard deviations (SDs), while non-normally distributed variables are presented as medians (interquartile ranges) and log-transformed for normal distribution before statistical analysis. Categorical data are presented as absolute numbers (percentages). Intergroup differences were evaluated using Student’s t-test or the Mann-Whitney test for continuous variables and the χ² test for categorical variables. Group comparisons were performed using ANOVA or the Bonferroni test for multiple comparisons. Stepwise linear regression was applied to explore the relationship between changes in echocardiographic parameters and baseline echocardiographic values, alongside various baseline clinical risk factors such as age, gender, hypertension duration, BMI, blood pressure, heart rate, smoking status, LDL-C, HbA1c, SCr, ACTH, 24h-UFC, medication type and quantity, and changes in BMI, blood pressure, and heart rate. All statistical analyses were conducted using SPSS for Windows (Version 23.0), with a two-sided P-value <0.05 considered statistically significant.

## 3. Results

### 3.1 Clinical characteristics of the entire cohort

The clinical characteristics and echocardiographic parameters at baseline and follow-up, along with their longitudinal changes, are summarized in **Table 1**. The mean age of the cohort was 44.68±12.20 years, with 29.8% being female. The mean follow-up period was 28.54±14.21 months. During this period, significant reductions were observed in BMI, systolic and diastolic blood pressure, and heart rate. The proportions of antihypertensive drug usage, including ACEI/ARB, ARNI, CCB, diuretics, spironolactone, beta-blockers, and alpha-blockers, remained consistent from baseline to the end of the study.

**Table 1:**
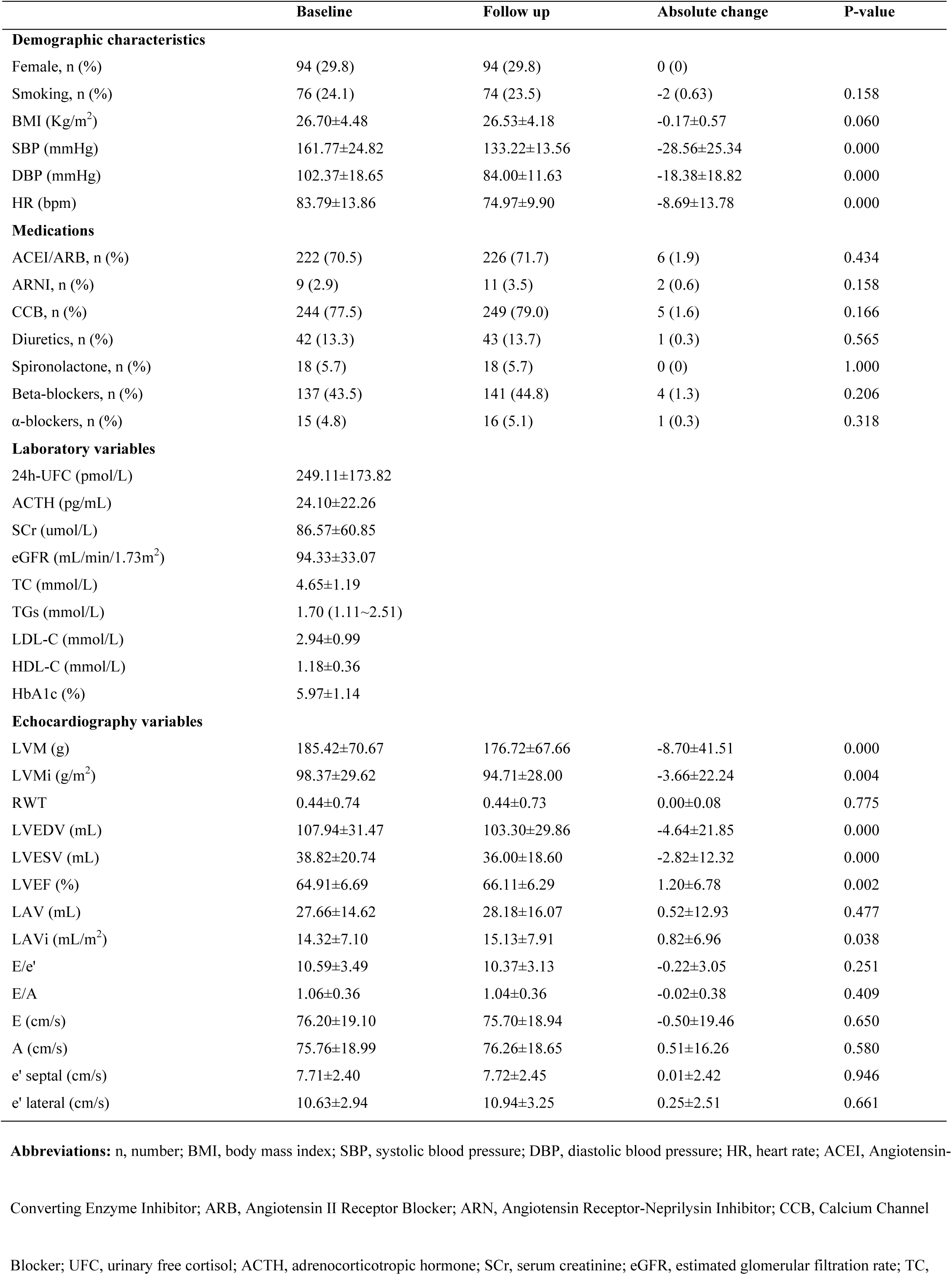
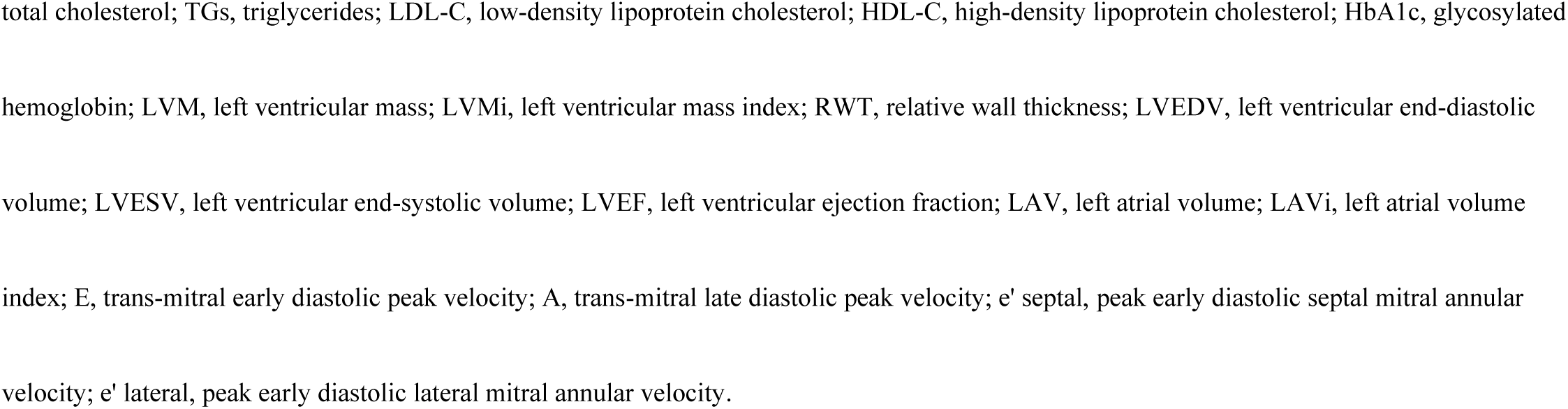
Clinical characteristics and echocardiography parameters of patients at baseline and follow-up.

|  | Baseline | Follow up | Absolute change | P-value |
| --- | --- | --- | --- | --- |
| **Demographic characteristics** |  |  |  |  |
| Female, n (%) | 94 (29.8) | 94 (29.8) | 0 (0) |  |
| Smoking, n (%) | 76 (24.1) | 74 (23.5) | -2 (0.63) | 0.158 |
| BMI (Kg/m<sup>2</sup>) | 26.70±4.48 | 26.53±4.18 | -0.17±0.57 | 0.060 |
| SBP (mmHg) | 161.77±24.82 | 133.22±13.56 | -28.56±25.34 | 0.000 |
| DBP (mmHg) | 102.37±18.65 | 84.00±11.63 | -18.38±18.82 | 0.000 |
| HR (bpm) | 83.79±13.86 | 74.97±9.90 | -8.69±13.78 | 0.000 |
| **Medications** |  |  |  |  |
| ACEI/ARB, n (%) | 222 (70.5) | 226 (71.7) | 6 (1.9) | 0.434 |
| ARNI, n (%) | 9 (2.9) | 11 (3.5) | 2 (0.6) | 0.158 |
| CCB, n (%) | 244 (77.5) | 249 (79.0) | 5 (1.6) | 0.166 |
| Diuretics, n (%) | 42 (13.3) | 43 (13.7) | 1 (0.3) | 0.565 |
| Spironolactone, n (%) | 18 (5.7) | 18 (5.7) | 0 (0) | 1.000 |
| Beta-blockers, n (%) | 137 (43.5) | 141 (44.8) | 4 (1.3) | 0.206 |
| α-blockers, n (%) | 15 (4.8) | 16 (5.1) | 1 (0.3) | 0.318 |
| **Laboratory variables** |  |  |  |  |
| 24h-UFC (pmol/L) | 249.11±173.82 |  |  |  |
| ACTH (pg/mL) | 24.10±22.26 |  |  |  |
| SCr (umol/L) | 86.57±60.85 |  |  |  |
| eGFR (mL/min/1.73m<sup>2</sup>) | 94.33±33.07 |  |  |  |
| TC (mmol/L) | 4.65±1.19 |  |  |  |
| TGs (mmol/L) | 1.70 (1.11~2.51) |  |  |  |
| LDL-C (mmol/L) | 2.94±0.99 |  |  |  |
| HDL-C (mmol/L) | 1.18±0.36 |  |  |  |
| HbA1c (%) | 5.97±1.14 |  |  |  |
| **Echocardiography variables** |  |  |  |  |
| LVM (g) | 185.42±70.67 | 176.72±67.66 | -8.70±41.51 | 0.000 |
| LVMi (g/m<sup>2</sup>) | 98.37±29.62 | 94.71±28.00 | -3.66±22.24 | 0.004 |
| RWT | 0.44±0.74 | 0.44±0.73 | 0.00±0.08 | 0.775 |
| LVEDV (mL) | 107.94±31.47 | 103.30±29.86 | -4.64±21.85 | 0.000 |
| LVESV (mL) | 38.82±20.74 | 36.00±18.60 | -2.82±12.32 | 0.000 |
| LVEF (%) | 64.91±6.69 | 66.11±6.29 | 1.20±6.78 | 0.002 |
| LAV (mL) | 27.66±14.62 | 28.18±16.07 | 0.52±12.93 | 0.477 |
| LAVi (mL/m<sup>2</sup>) | 14.32±7.10 | 15.13±7.91 | 0.82±6.96 | 0.038 |
| E/e' | 10.59±3.49 | 10.37±3.13 | -0.22±3.05 | 0.251 |
| E/A | 1.06±0.36 | 1.04±0.36 | -0.02±0.38 | 0.409 |
| E (cm/s) | 76.20±19.10 | 75.70±18.94 | -0.50±19.46 | 0.650 |
| A (cm/s) | 75.76±18.99 | 76.26±18.65 | 0.51±16.26 | 0.580 |
| e' septal (cm/s) | 7.71±2.40 | 7.72±2.45 | 0.01±2.42 | 0.946 |
| e' lateral (cm/s) | 10.63±2.94 | 10.94±3.25 | 0.25±2.51 | 0.661 |

**Abbreviations:** n, number; BMI, body mass index; SBP, systolic blood pressure; DBP, diastolic blood pressure; HR, heart rate; ACEI, Angiotensin-Converting Enzyme Inhibitor; ARB, Angiotensin II Receptor Blocker; ARN, Angiotensin Receptor-Neprilysin Inhibitor; CCB, Calcium Channel Blocker; UFC, urinary free cortisol; ACTH, adrenocorticotrophic hormone; SCr, serum creatinine; eGFR, estimated glomerular filtration rate; TC,
total cholesterol; TGs, triglycerides; LDL-C, low-density lipoprotein cholesterol; HDL-C, high-density lipoprotein cholesterol; HbA1c, glycosylated hemoglobin; LVM, left ventricular mass; LVMi, left ventricular mass index; RWT, relative wall thickness; LVEDV, left ventricular end-diastolic volume; LVESV, left ventricular end-systolic volume; LVEF, left ventricular ejection fraction; LAV, left atrial volume; LAVi, left atrial volume index; E, trans-mitral early diastolic peak velocity; A, trans-mitral late diastolic peak velocity; e' septal, peak early diastolic septal mitral annular velocity; e' lateral, peak early diastolic lateral mitral annular velocity.

**Table 2:**
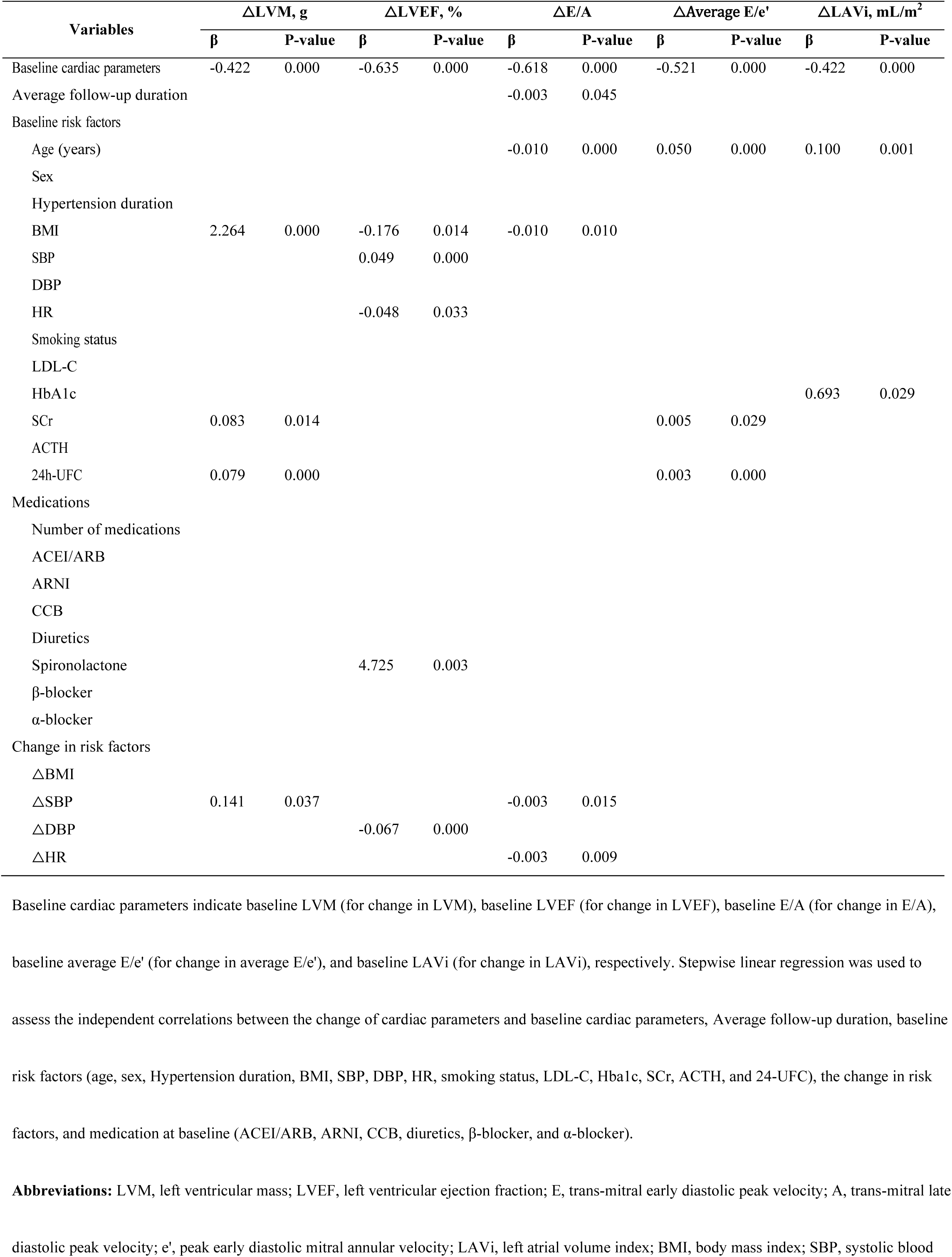
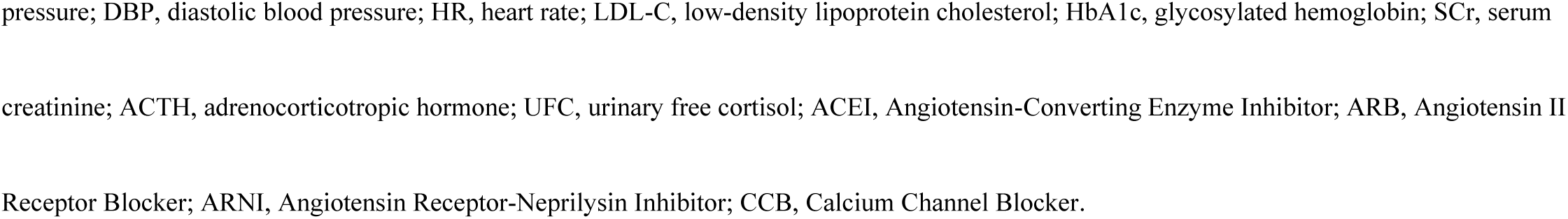
Factors related to changes in cardiac structure and function parameters.

| Variables | ΔLVM, g |  | ΔLVEF, % |  | ΔE/A |  | ΔAverage E/e' |  | ΔLAVi, mL/m<sup>2</sup> |  |
| --- | --- | --- | --- | --- | --- | --- | --- | --- | --- | --- |
|  | β | P-value | β | P-value | β | P-value | β | P-value | β | P-value |
| Baseline cardiac parameters | -0.422 | 0.000 | -0.635 | 0.000 | -0.618 | 0.000 | -0.521 | 0.000 | -0.422 | 0.000 |
| Average follow-up duration |  |  |  |  | -0.003 | 0.045 |  |  |  |  |
| Baseline risk factors |  |  |  |  |  |  |  |  |  |  |
| Age (years) |  |  |  |  | -0.010 | 0.000 | 0.050 | 0.000 | 0.100 | 0.001 |
| Sex |  |  |  |  |  |  |  |  |  |  |
| Hypertension duration |  |  |  |  |  |  |  |  |  |  |
| BMI | 2.264 | 0.000 | -0.176 | 0.014 | -0.010 | 0.010 |  |  |  |  |
| SBP |  |  | 0.049 | 0.000 |  |  |  |  |  |  |
| DBP |  |  |  |  |  |  |  |  |  |  |
| HR |  |  | -0.048 | 0.033 |  |  |  |  |  |  |
| Smoking status |  |  |  |  |  |  |  |  |  |  |
| LDL-C |  |  |  |  |  |  |  |  |  |  |
| HbA1c |  |  |  |  |  |  |  |  | 0.693 | 0.029 |
| SCr | 0.083 | 0.014 |  |  |  |  | 0.005 | 0.029 |  |  |
| ACTH |  |  |  |  |  |  |  |  |  |  |
| 24h-UFC | 0.079 | 0.000 |  |  |  |  | 0.003 | 0.000 |  |  |
| Medications |  |  |  |  |  |  |  |  |  |  |
| Number of medications |  |  |  |  |  |  |  |  |  |  |
| ACEI/ARB |  |  |  |  |  |  |  |  |  |  |
| ARNI |  |  |  |  |  |  |  |  |  |  |
| CCB |  |  |  |  |  |  |  |  |  |  |
| Diuretics |  |  |  |  |  |  |  |  |  |  |
| Spironolactone |  |  | 4.725 | 0.003 |  |  |  |  |  |  |
| β-blocker |  |  |  |  |  |  |  |  |  |  |
| α-blocker |  |  |  |  |  |  |  |  |  |  |
| Change in risk factors |  |  |  |  |  |  |  |  |  |  |
| ΔBMI |  |  |  |  |  |  |  |  |  |  |
| ΔSBP | 0.141 | 0.037 |  |  | -0.003 | 0.015 |  |  |  |  |
| ΔDBP |  |  | -0.067 | 0.000 |  |  |  |  |  |  |
| ΔHR |  |  |  |  | -0.003 | 0.009 |  |  |  |  |

Baseline cardiac parameters indicate baseline LVM (for change in LVM), baseline LVEF (for change in LVEF), baseline E/A (for change in E/A), baseline average E/e' (for change in average E/e'), and baseline LAVi (for change in LAVi), respectively. Stepwise linear regression was used to assess the independent correlations between the change of cardiac parameters and baseline cardiac parameters, Average follow-up duration, baseline risk factors (age, sex, Hypertension duration, BMI, SBP, DBP, HR, smoking status, LDL-C, Hba1c, SCr, ACTH, and 24-UFC), the change in risk factors, and medication at baseline (ACEI/ARB, ARNI, CCB, diuretics, β-blocker, and α-blocker).
**Abbreviations:** LVM, left ventricular mass; LVEF, left ventricular ejection fraction; E, trans-mitral early diastolic peak velocity; A, trans-mitral late diastolic peak velocity; e', peak early diastolic mitral annular velocity; LAVi, left atrial volume index; BMI, body mass index; SBP, systolic blood
pressure; DBP, diastolic blood pressure; HR, heart rate; LDL-C, low-density lipoprotein cholesterol; HbA1c, glycosylated hemoglobin; SCr, serum creatinine; ACTH, adrenocorticotrophic hormone; UFC, urinary free cortisol; ACEI, Angiotensin-Converting Enzyme Inhibitor; ARB, Angiotensin II Receptor Blocker; ARNI, Angiotensin Receptor-Neprilysin Inhibitor; CCB, Calcium Channel Blocker.

**Table 3:**
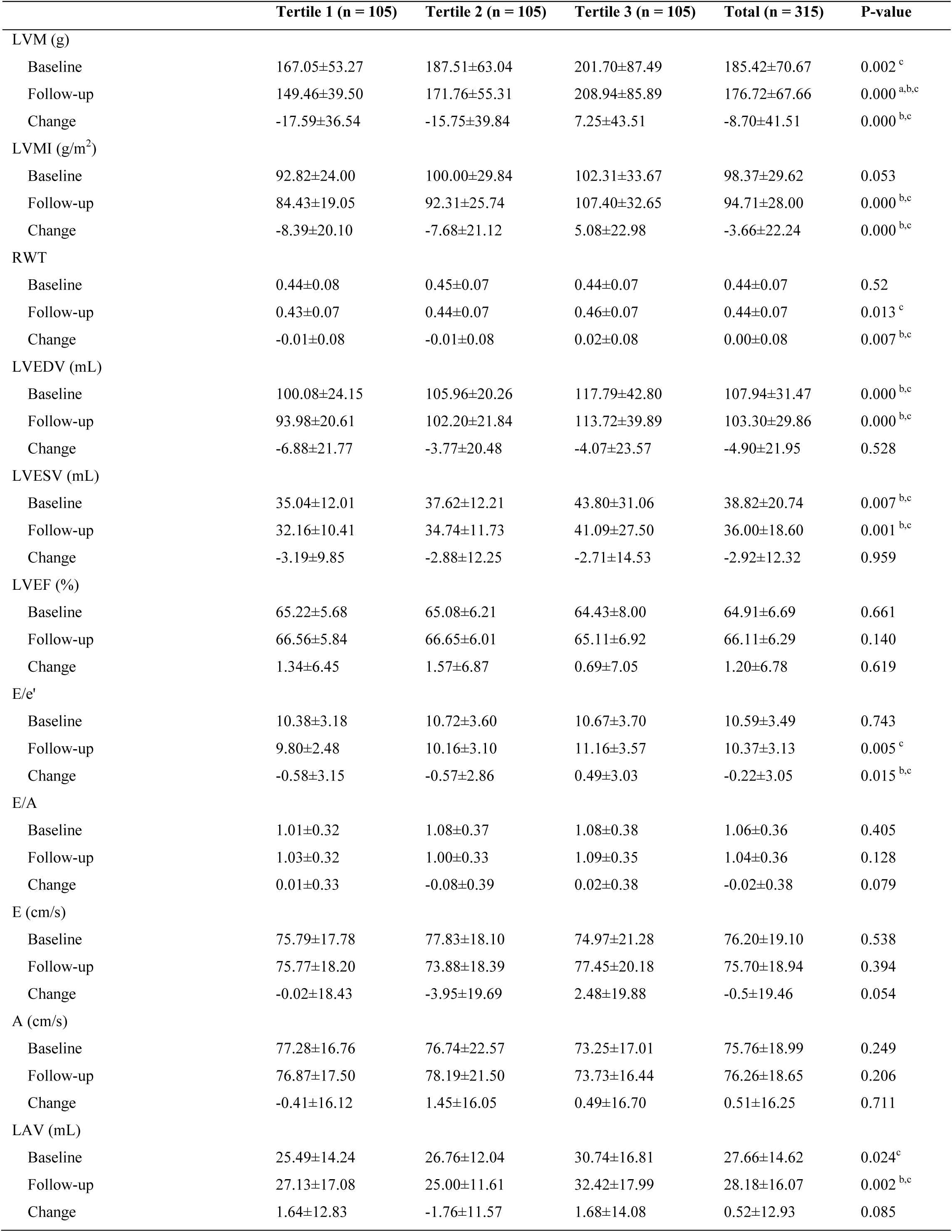
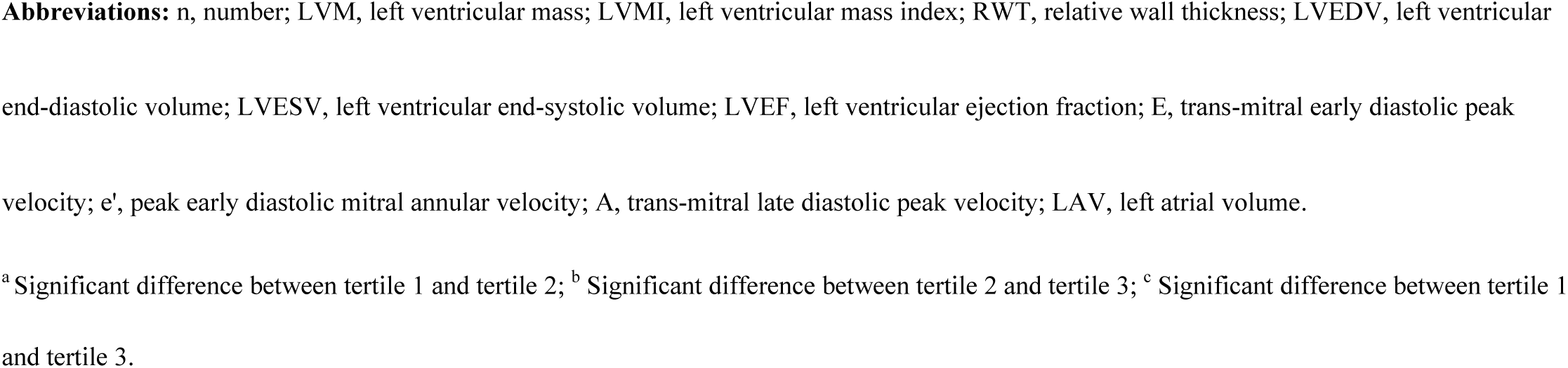
Echocardiographic parameters of patients grouped according to tertiles of 24h-UFC.

|  | Tertile 1 (n = 105) | Tertile 2 (n = 105) | Tertile 3 (n = 105) | Total (n = 315) | P-value |
| --- | --- | --- | --- | --- | --- |
| LVM (g) |  |  |  |  |  |
| Baseline | 167.05±53.27 | 187.51±63.04 | 201.70±87.49 | 185.42±70.67 | 0.002<sup>c</sup> |
| Follow-up | 149.46±39.50 | 171.76±55.31 | 208.94±85.89 | 176.72±67.66 | 0.000<sup>a,b,c</sup> |
| Change | -17.59±36.54 | -15.75±39.84 | 7.25±43.51 | -8.70±41.51 | 0.000<sup>b,c</sup> |
| LVMI (g/m<sup>2</sup>) |  |  |  |  |  |
| Baseline | 92.82±24.00 | 100.00±29.84 | 102.31±33.67 | 98.37±29.62 | 0.053 |
| Follow-up | 84.43±19.05 | 92.31±25.74 | 107.40±32.65 | 94.71±28.00 | 0.000<sup>b,c</sup> |
| Change | -8.39±20.10 | -7.68±21.12 | 5.08±22.98 | -3.66±22.24 | 0.000<sup>b,c</sup> |
| RWT |  |  |  |  |  |
| Baseline | 0.44±0.08 | 0.45±0.07 | 0.44±0.07 | 0.44±0.07 | 0.52 |
| Follow-up | 0.43±0.07 | 0.44±0.07 | 0.46±0.07 | 0.44±0.07 | 0.013<sup>c</sup> |
| Change | -0.01±0.08 | -0.01±0.08 | 0.02±0.08 | 0.00±0.08 | 0.007<sup>b,c</sup> |
| LVEDV (mL) |  |  |  |  |  |
| Baseline | 100.08±24.15 | 105.96±20.26 | 117.79±42.80 | 107.94±31.47 | 0.000<sup>b,c</sup> |
| Follow-up | 93.98±20.61 | 102.20±21.84 | 113.72±39.89 | 103.30±29.86 | 0.000<sup>b,c</sup> |
| Change | -6.88±21.77 | -3.77±20.48 | -4.07±23.57 | -4.90±21.95 | 0.528 |
| LVESV (mL) |  |  |  |  |  |
| Baseline | 35.04±12.01 | 37.62±12.21 | 43.80±31.06 | 38.82±20.74 | 0.007<sup>b,c</sup> |
| Follow-up | 32.16±10.41 | 34.74±11.73 | 41.09±27.50 | 36.00±18.60 | 0.001<sup>b,c</sup> |
| Change | -3.19±9.85 | -2.88±12.25 | -2.71±14.53 | -2.92±12.32 | 0.959 |
| LVEF (%) |  |  |  |  |  |
| Baseline | 65.22±5.68 | 65.08±6.21 | 64.43±8.00 | 64.91±6.69 | 0.661 |
| Follow-up | 66.56±5.84 | 66.65±6.01 | 65.11±6.92 | 66.11±6.29 | 0.140 |
| Change | 1.34±6.45 | 1.57±6.87 | 0.69±7.05 | 1.20±6.78 | 0.619 |
| E/e' |  |  |  |  |  |
| Baseline | 10.38±3.18 | 10.72±3.60 | 10.67±3.70 | 10.59±3.49 | 0.743 |
| Follow-up | 9.80±2.48 | 10.16±3.10 | 11.16±3.57 | 10.37±3.13 | 0.005<sup>c</sup> |
| Change | -0.58±3.15 | -0.57±2.86 | 0.49±3.03 | -0.22±3.05 | 0.015<sup>b,c</sup> |
| E/A |  |  |  |  |  |
| Baseline | 1.01±0.32 | 1.08±0.37 | 1.08±0.38 | 1.06±0.36 | 0.405 |
| Follow-up | 1.03±0.32 | 1.00±0.33 | 1.09±0.35 | 1.04±0.36 | 0.128 |
| Change | 0.01±0.33 | -0.08±0.39 | 0.02±0.38 | -0.02±0.38 | 0.079 |
| E (cm/s) |  |  |  |  |  |
| Baseline | 75.79±17.78 | 77.83±18.10 | 74.97±21.28 | 76.20±19.10 | 0.538 |
| Follow-up | 75.77±18.20 | 73.88±18.39 | 77.45±20.18 | 75.70±18.94 | 0.394 |
| Change | -0.02±18.43 | -3.95±19.69 | 2.48±19.88 | -0.5±19.46 | 0.054 |
| A (cm/s) |  |  |  |  |  |
| Baseline | 77.28±16.76 | 76.74±22.57 | 73.25±17.01 | 75.76±18.99 | 0.249 |
| Follow-up | 76.87±17.50 | 78.19±21.50 | 73.73±16.44 | 76.26±18.65 | 0.206 |
| Change | -0.41±16.12 | 1.45±16.05 | 0.49±16.70 | 0.51±16.25 | 0.711 |
| LAV (mL) |  |  |  |  |  |
| Baseline | 25.49±14.24 | 26.76±12.04 | 30.74±16.81 | 27.66±14.62 | 0.024<sup>c</sup> |
| Follow-up | 27.13±17.08 | 25.00±11.61 | 32.42±17.99 | 28.18±16.07 | 0.002<sup>b,c</sup> |
| Change | 1.64±12.83 | -1.76±11.57 | 1.68±14.08 | 0.52±12.93 | 0.085 |

**Abbreviations:** n, number; LVM, left ventricular mass; LVMI, left ventricular mass index; RWT, relative wall thickness; LVEDV, left ventricular end-diastolic volume; LVESV, left ventricular end-systolic volume; LVEF, left ventricular ejection fraction; E, trans-mitral early diastolic peak velocity; e', peak early diastolic mitral annular velocity; A, trans-mitral late diastolic peak velocity; LAV, left atrial volume.
<sup>a</sup> Significant difference between tertile 1 and tertile 2; <sup>b</sup> Significant difference between tertile 2 and tertile 3; <sup>c</sup> Significant difference between tertile 1 and tertile 3.

### 3.2 Echocardiographic parameters of the entire cohort

The cohort demonstrated a significant reduction in LV structure parameters, including LVMi, LVESV, and LVEDV at follow-up, while RWT and LAVi remained similar. The prevalence of LVH decreased from 26.4% at baseline to 23.8% at follow-up, though this reduction was not statistically significant (**Figure 2**). Additionally, the mean LVEF significantly increased from 64.91 ± 6.69% to 66.11 ± 6.29% (p=0.002). However, there was no significant change in the proportion of patients with diastolic dysfunction during the follow-up period (**Figure 3**).

**Figure 2.**
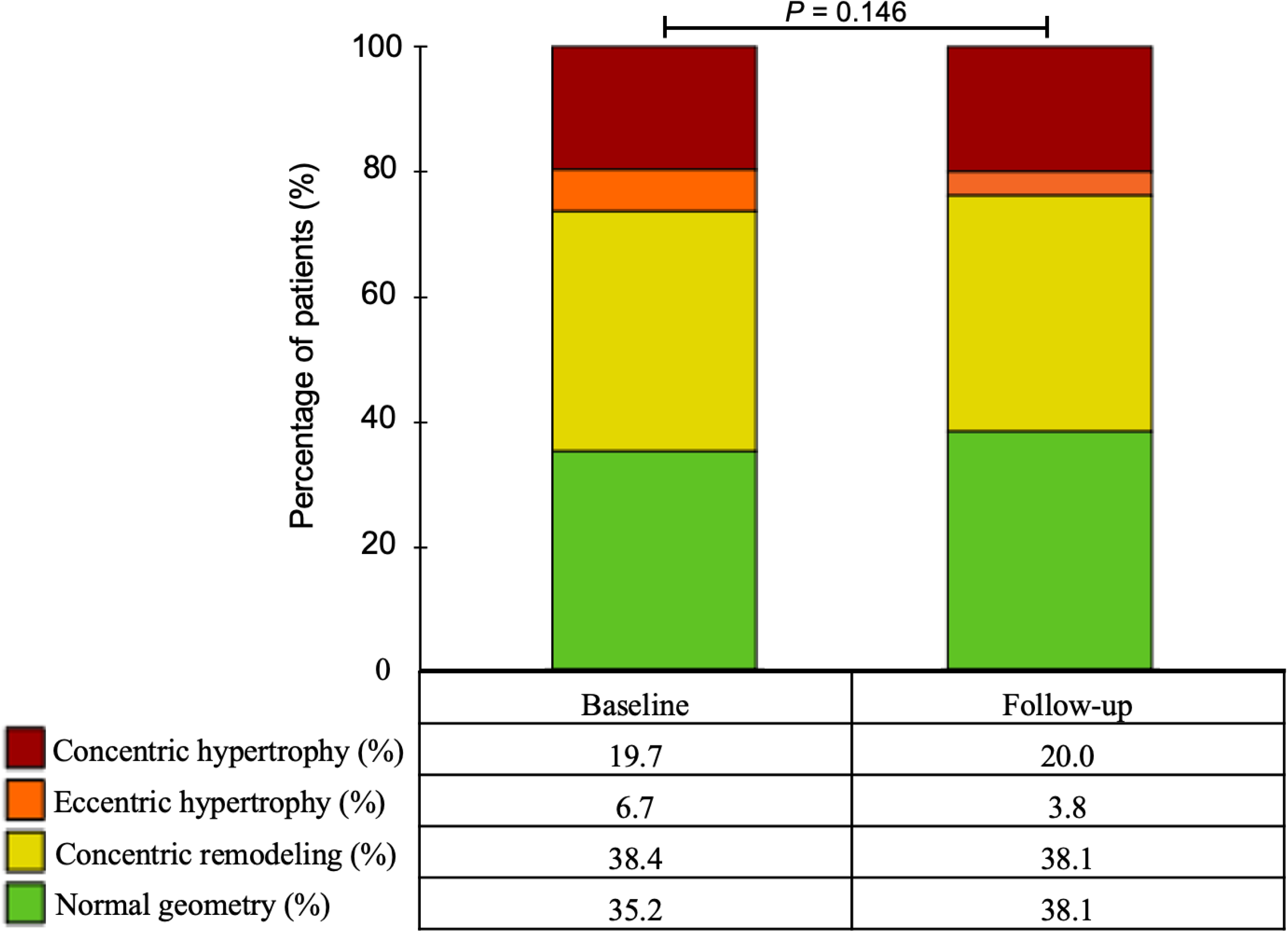
Longitudinal changes in the proportion of LVH types in the entire cohort.

**Figure 3.**
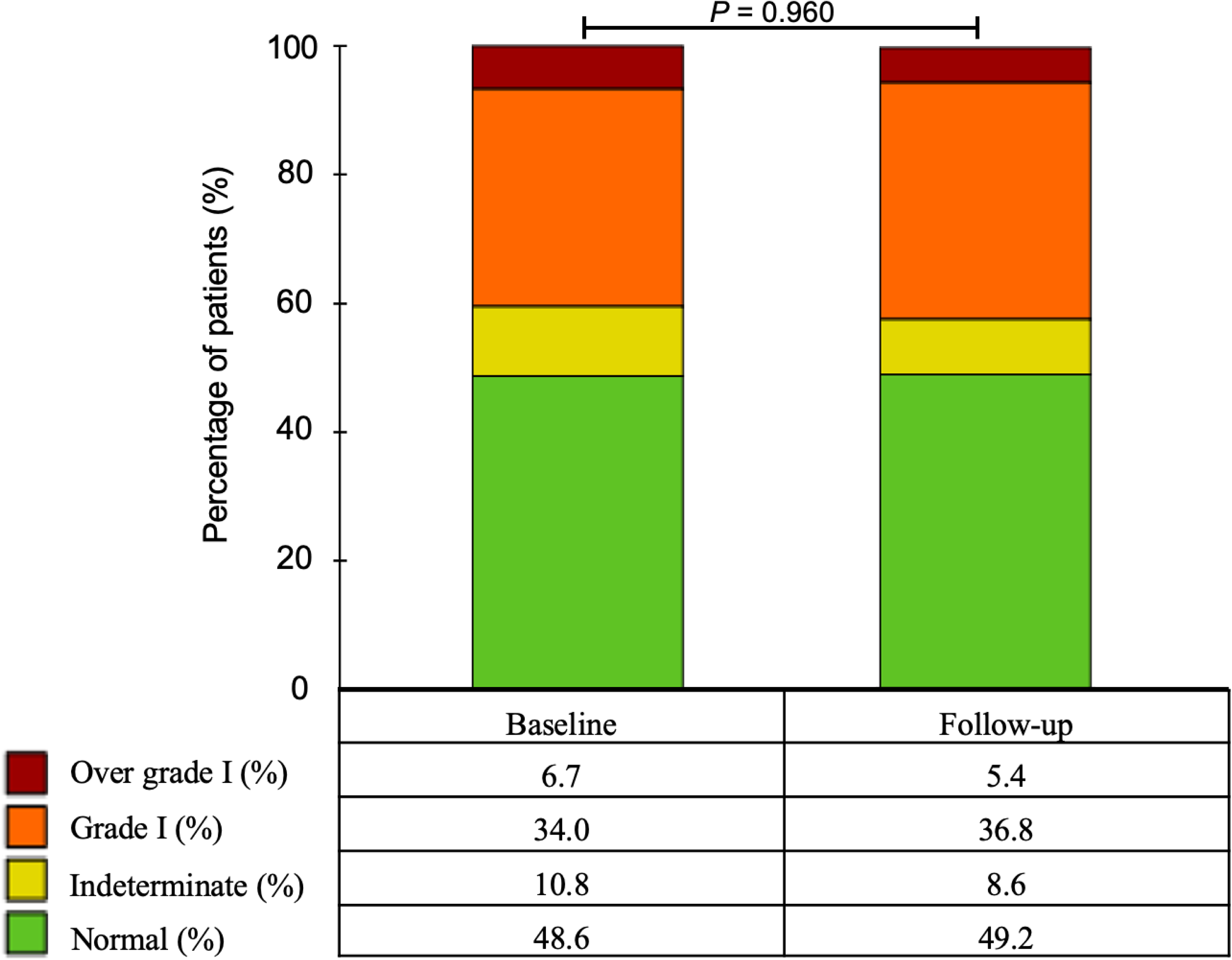
Longitudinal changes in the proportion of LV diastolic dysfunction in the entire cohort.

**Figure 4.**
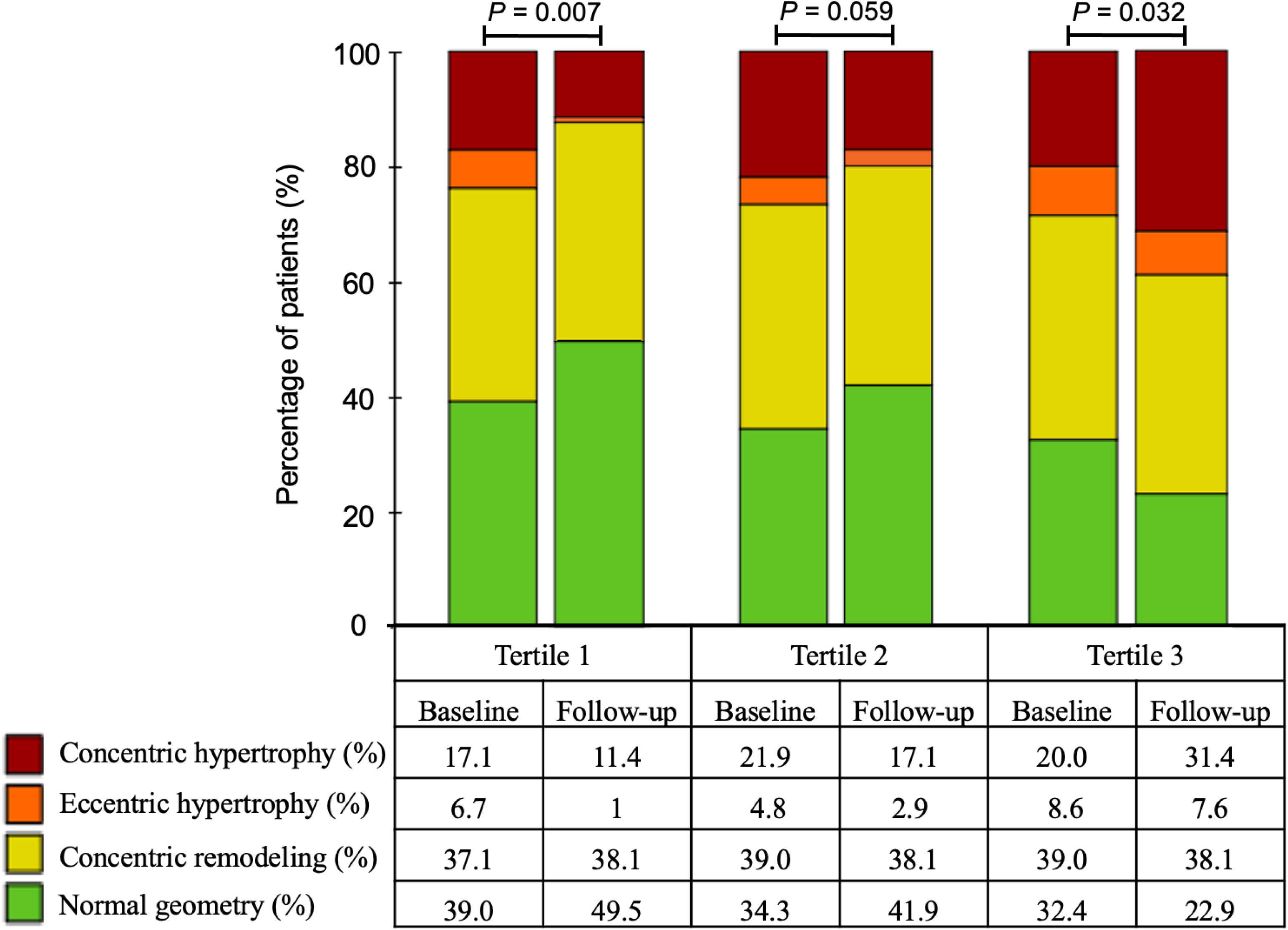
Longitudinal changes in the proportion of LVH types of patients grouped according to tertiles of 24h-UFC.

**Figure 5.**
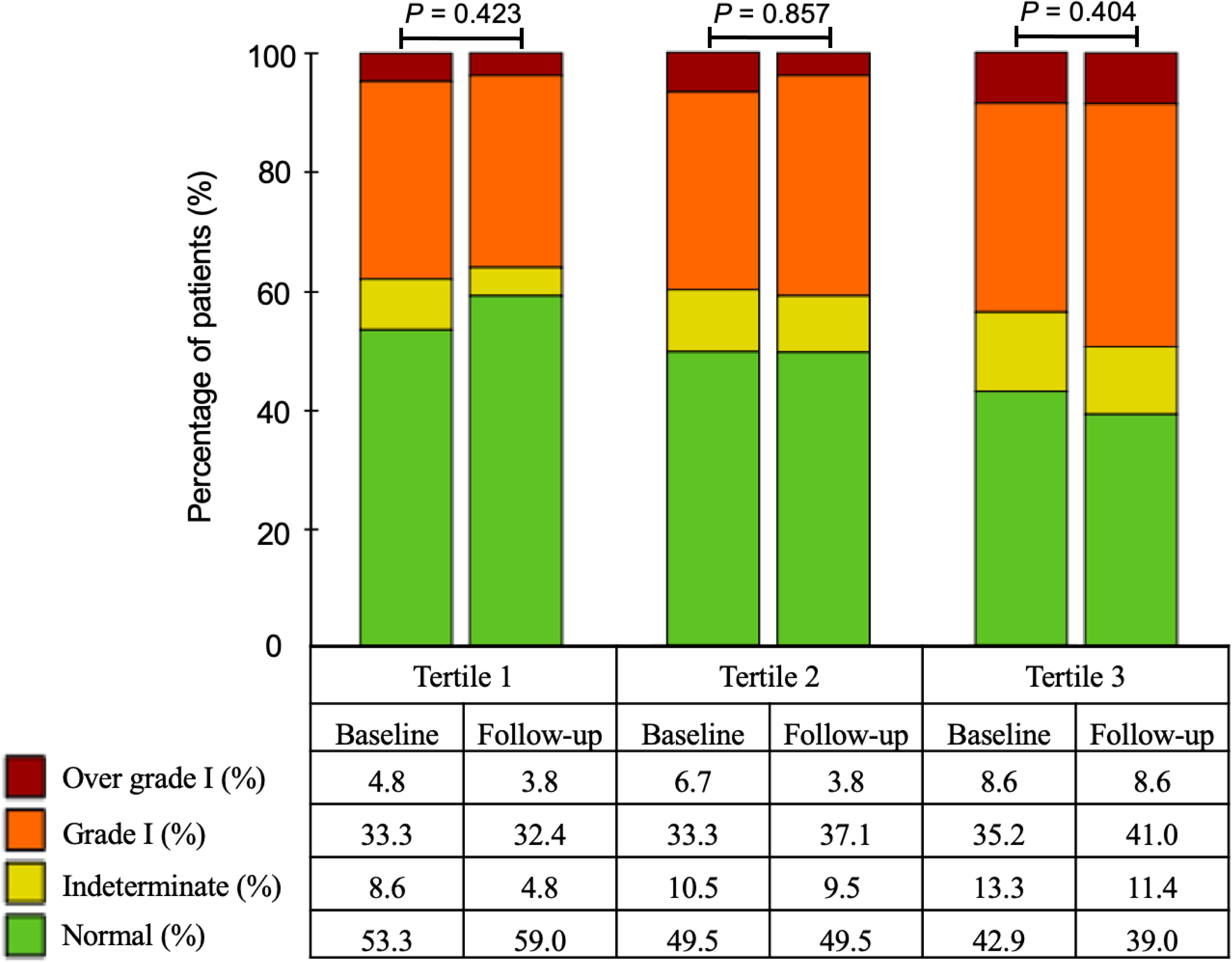
Longitudinal changes in the proportion of LV diastolic dysfunction of patients grouped according to tertiles of 24h-UFC.

### 3.3 Factors associated with changes in LV structure and function

## 4. Discussion

To the best of our knowledge, this is the first prospective cohort study to examine the relationship between longitudinal changes in cardiac structure/function and 24h-UFC in essential hypertension. The most significant finding of our study is that baseline 24h-UFC levels are significantly associated with adverse LV remodeling in these patients, even after adjusting for various confounding factors.

Previous studies in patients with Cushing’s syndrome have shown that excessive cortisol levels can independently induce abnormalities in LV structure and function, irrespective of blood pressure changes^17–19^. Similar findings have been reported in small-sample cross-sectional studies in patients with essential hypertension.^11,20^ However, the cross-sectional nature of these studies does not allow for definitive conclusions regarding a causal relationship between cortisol levels and LV changes. Consequently, we designed the current prospective cohort study to examine the relationship between baseline cortisol levels and longitudinal changes in LV structure and function. In this study, we used 24h-UFC as a measure of circulating free cortisol levels. This method provides a direct assessment of free cortisol in circulation, independent of corticosteroid-binding globulin (CBG), and minimizes the impact of cortisol’s circadian rhythm, thus offering a more accurate reflection of tissue and organ exposure to active cortisol. ^21,22^ Our analysis revealed that changes in LVM (ΔLVM) were independently associated with baseline 24h-UFC levels. We further investigated changes in LV geometry from baseline to the end of follow-up among patients with different 24h-UFC levels. Notably, patients in the highest 24h-UFC tertile exhibited a higher proportion of LVH at baseline and a further increase during follow-up, particularly in concentric hypertrophy, even after antihypertensive treatment. In contrast, patients with lower and moderate cortisol levels showed a reduction in the proportion of LVH following antihypertensive treatment. This difference cannot be fully explained by blood pressure control, as there were no significant differences in changes in systolic and diastolic blood pressure (ΔSBP and ΔDBP) among the three groups. Previous studies in Cushing’s syndrome patients have also demonstrated that the reversibility of LV structural changes is not related to post-treatment blood pressure changes.^9,18^ Beyond hypertension, chronic exposure to high cortisol levels is associated with other metabolic abnormalities, including central obesity, insulin resistance, hyperglycemia, and dyslipidemia, all of which may contribute to LV hypertrophy, fibrosis, and fat deposition.^23^ In addition to these metabolic factors, the direct effects of cortisol on the cardiovascular system must be considered. The cardiovascular system is a primary target of cortisol, but the pathophysiological mechanisms underlying its direct effects remain complex and not fully elucidated.^24^ Cortisol exerts its biological effects primarily through binding to glucocorticoid receptors (GR) and mineralocorticoid receptors (MR), with these effects modulated by 11β-hydroxysteroid dehydrogenase type 1 (HSD1) and type 2 (HSD2).^25,26^ 11β-HSD1 converts inactive cortisone to active cortisol, increasing both circulating and local active cortisol concentrations, while 11β-HSD2 inactivates cortisol.^27–29^ Therefore, the effects of cortisol on target cells are closely linked to the balance between 11β-HSD1 and 11β-HSD2 both in circulation and within cells. At physiological levels, cortisol helps maintain cardiac performance by regulating the life cycle of cardiomyocytes, including growth, differentiation, metabolism, and apoptosis.^30^ However, excessive levels of glucocorticoids can lead to overactivation of GR, promoting cardiomyocyte hypertrophy.^31^ Studies have shown that exposure of rat cardiomyocytes to dexamethasone results in significant increases in cell size and elevated expression levels of hypertrophy markers such as atrial natriuretic factor (ANF), β-myosin heavy chain (β-MHC), and skeletal muscle α-actin (α-SKA), effects that were mitigated by GR antagonists or GR knockdown.^32^ Additionally, cortisol can directly activate MR in cardiomyocytes, producing aldosterone-like effects and promoting the progression of LVH.^33,34^ Under physiological conditions, circulating cortisol levels are over 100 times higher than aldosterone levels, with greater affinity for MRs than aldosterone.^35^ Furthermore, cortisol has been shown to enhance the effects of noradrenaline and Angiotensin II, thereby promoting LVH by enhancing the sympathetic nervous system and the renin-angiotensin-aldosterone system (RAAS). In addition to its actions on circulating RAAS, cortisol can also activate myocardial tissue RAAS via paracrine pathways, leading to cardiomyocyte proliferation and hypertrophy.^36^

Our study also demonstrates that baseline 24h-UFC levels are independently positively correlated with changes in the E/e’ ratio (ΔE/e’). We, therefore, further investigated alterations in LV diastolic function from baseline to follow-up among patients with varying 24h-UFC levels. The results indicated no significant changes in the proportion of patients with LV diastolic dysfunction across the three groups from baseline to follow-up. However, in contrast to the groups with lower and moderate cortisol levels, the group with the highest cortisol levels exhibited a trend towards a further increase in the proportion of patients with LV diastolic dysfunction, despite effective blood pressure control. Previous research has shown that patients with Cushing’s syndrome, characterized by excessive cortisol secretion, have a higher prevalence of LV diastolic dysfunction compared to those with essential hypertension.^37^ Furthermore, a cross-sectional study in patients with essential hypertension revealed that the prevalence of LV diastolic dysfunction increases with higher DST cortisol levels.^11^ Recent studies in patients with type 2 diabetes have also demonstrated a significant positive correlation between plasma cortisol concentration and the E/e’ ratio.^38^ The relationship between cortisol levels and LV diastolic dysfunction is not entirely attributable to LVH. Research has indicated that, compared to healthy subjects and patients with essential hypertension, untreated patients with Cushing’s syndrome exhibit significantly increased myocardial fibrosis and more severe LV diastolic dysfunction.^8^ Following treatment for Cushing’s syndrome, both myocardial fibrosis and LV diastolic function can improve, suggesting that excessive cortisol may promote myocardial fibrosis, thereby contributing to the development of LV diastolic dysfunction. The absence of statistically significant changes in the proportion of patients with LV diastolic dysfunction at the end of follow-up in our study may be due to the relatively small sample size and the insufficient duration of follow-up.

## 5. Limitations

The limitations of this study should be acknowledged. The relatively small sample size and the considerable variability in the intervals between ultrasound follow-ups among individuals somewhat weaken the strength of the study’s conclusions. Moreover, assessing cortisol exposure based on a single 24h-UFC measurement may not be fully reliable due to daily fluctuations in 24h-UFC levels.^39^ Additionally, the study measured only baseline 24h-UFC levels, without tracking dynamic changes over time. Future larger-scale studies are needed to clarify the independent effects of cortisol levels on changes in left ventricular structure and function in patients with essential hypertension.

## 6. Clinical implications

In patients with essential hypertension, a subset experiences further progression of LV remodeling and diastolic dysfunction despite antihypertensive treatment. Identifying factors beyond elevated blood pressure that contribute to LV remodeling is crucial for developing therapies that effectively prevent or reverse this remodeling. Our study suggests an independent association between 24h-UFC levels and the progression of LV remodeling in hypertensive patients. Even with equally effective blood pressure control, patients with higher 24h-UFC levels exhibit more pronounced progression of LV remodeling. These findings highlight new potential avenues for the prevention and treatment of LV remodeling in hypertensive patients. Future studies should explore methods to reduce excess cortisol to prevent the development of LV remodeling.

## 7. Conclusions

Elevated 24h-UFC levels are independently associated with adverse changes in LV structure and diastolic function in patients with essential hypertension. These findings suggest that cortisol plays a significant role in promoting LV hypertrophy and diastolic dysfunction, beyond the effects of blood pressure. Future research should explore interventions targeting cortisol reduction to prevent or reverse cardiac remodeling in hypertensive patients with elevated cortisol levels.

## Data Availability

The data supporting this study are available from the corresponding author upon reasonable request.

## Acknowledgments

The authors have no conflicts of interest to disclose. All authors critically reviewed and contributed to the manuscript’s intellectual content. KHY and JCX were responsible for the study’s conception. GZC carried out the initial data preparation. Statistical analyses were conducted by GZC, with support from KHY and JCX, and additional input from JYH, RW, CC and QSL. GZC and KHY drafted multiple versions of the manuscript. KHY and JCX served as the guarantors of this work, having full access to all the data and assuming responsibility for the integrity of the data and the accuracy of the analysis. All authors have reviewed and approved the final version of the manuscript.

